# Improving cardiovascular risk prediction beyond pooled cohort equations: a prospective cohort of 304,356 participants

**DOI:** 10.1101/2023.01.09.23284368

**Authors:** Joshua Elliott, Barbara Bodinier, Matthew Whitaker, Ioanna Tzoulaki, Paul Elliott, Marc Chadeau-Hyam

**Author notes:** **Corresponding Authors:** Marc Chadeau-Hyam, Department of Epidemiology and Biostatistics, School of Public Health, St Mary’s Campus, Norfolk Place, Medical School Building, 5^th^ Floor, Imperial College London, London W21PG, United Kingdom Paul Elliott, Department of Epidemiology and Biostatistics, School of Public Health, St Mary’s Campus, Norfolk Place, Medical School Building, 1^st^ Floor, Imperial College London, London W21PG, United Kingdom ( k). Joint first authors (Joshua Elliott, Barbara Bodinier). Joint last authors (Paul Elliott, Marc Chadeau-Hyam).

## Abstract

**Background:** Pooled Cohort Equations (PCE) are used to predict cardiovascular disease (CVD) risk. Inclusion of other variables may improve risk prediction.

**Objective:** Identify variables improving CVD risk prediction beyond recalibrated PCE.

**Design:** Prospective cohort study; sex-stratified Cox survival models with LASSO stability selection to predict CVD in non-overlapping subsets: variable selection (40%), model training (30%) and testing (30%).

**Setting:** UK population.

**Participants:** UK Biobank: 121,724 and 182,632 healthy men and women, respectively, aged 38-73 years at baseline.

**Measurements:** Personal/family medical history; lifestyle factors; genetic, biochemical, hematological, and metabolomic blood markers. Outcomes were incident hospitalization or mortality from CVD.

**Results:** There were 11,899 (men) and 9,110 (women) incident CVD cases with median 12.1 years follow-up. Variables selected for both men and women were: age, albumin, antihypertensive medication, apolipoprotein B, atrial fibrillation, C-reactive protein, current smoker, cystatin C, family history of coronary artery disease, glycated hemoglobin, polygenic risk score (PRS) for CVD and systolic blood pressure. Also selected: apolipoprotein A1, lipoprotein(a), white blood cell count, deprivation index (men); triglycerides (women). C-statistics for recalibrated PCE were 0.67 [0.66-0.68] and 0.69 [0.68-0.70] in men and women, respectively, improving to 0.71 [0.70-0.72] and 0.72 [0.71-0.73] with LASSO stably selected variables. Categorical net reclassification improvement (7.5% risk threshold) versus PCE was 0.054 [0.038-0.070] (men) and 0.081 [0.063-0.099] (women). Addition of targeted metabolomic data to LASSO stability selection did not improve predictive accuracy.

**Limitations:** Analyses were done in a single population study and require external replication.

**Conclusion:** Additional personal/family medical history, blood-based markers and genetic information improve CVD risk prediction beyond PCE.

**Funding source:** National Institute for Health Research Academic Clinical Fellowship (JE); Medical Research Council studentship (BB); European Union H2020 (MC-H).

## Introduction

Cardiovascular disease (CVD) is the leading cause of morbidity and mortality worldwide. (1) Risk stratification via accurate prediction of future CVD risk is key to guiding effective early management and prevention. A systematic review identified 363 CVD prediction models that included more than 100 distinct variables; model size ranged from two to 80 variables, with seven on average. (2) The most commonly included variables were age, smoking, systolic blood pressure, history of diabetes, total cholesterol and high-density lipoprotein cholesterol. These predictors, along with ethnicity and antihypertensive use, are included in pooled cohort equations (PCE), which are used in the US to predict CVD risk. (3,4) In the UK, QRISK3 is used and incorporates additional variables. (5) Current US guidelines recommend lipid-lowering (statin) therapy for individuals with 10-year absolute risk of atherosclerotic CVD greater than 7.5%, as calculated by PCE, and low-density lipoprotein cholesterol levels higher than 70 mg/dL. (3)

Machine learning models have shown increasing promise in predicting incident coronary artery disease (CAD), with one recent example combining data from electronic health care records with a polygenic risk score (PRS) for CAD (6). Other recent studies have reported that PRS for CAD/CVD, when considered in isolation, yield a modest or non-significant improvement in predictive performance for CVD risk over traditional risk models. (7–11) It has also been suggested that metabolomic data may be predictive of incident CVD, although their clinical utility in risk prediction remains to be established (12–15). Here, we use the UK Biobank dataset to evaluate whether inclusion of a range of variables, spanning personal/family medical history, lifestyle factors, blood-based biochemical, metabolomic and genetic (PRS) markers could improve CVD risk prediction beyond PCE.

## Methods

### Study participants

UK Biobank recruited 502,536 volunteers aged 38–73 years between 2006 and 2010. Demographic and lifestyle factors, medical and surgical histories, standardized clinical measurements and blood samples were collected at baseline. A panel of laboratory tests was performed on stored serum and red blood cells as well as genotyping. (16) We excluded a total of 198,180 participants: 151,806 with prevalent CVD or missing data for any of the variables included in PCE or QRISK3 (9), 45,887 on lipid-lowering agents and a further 487 who had withdrawn consent, leaving 304,356 participants without prior CVD at baseline for the present analyses (121,724 men and 182,632 women, Figure 1). Among these, a subset of 27,873 men and 40,982 women also had data on nuclear magnetic resonance (NMR) metabolic biomarkers measured in baseline plasma samples.

**Figure 1.**
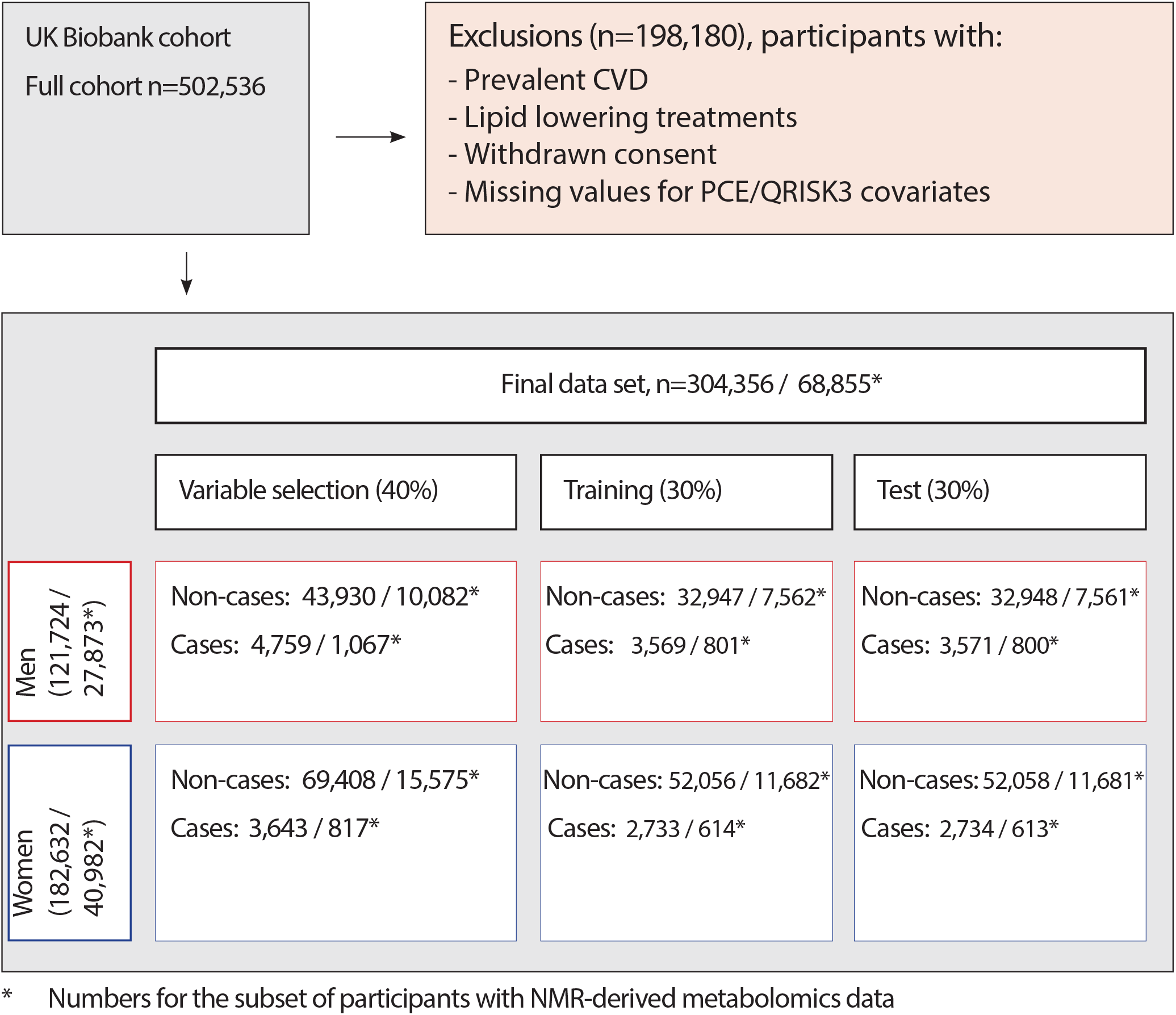
Study design and flowchart. Cases are participants with a cardiovascular disease event during follow-up including myocardial infarction and its sequelae, angina, non-hemorrhagic stroke and transient ischemic attack. Data were randomly split into three sex-stratified, non-overlapping sets: i) variable selection dataset (40%); ii) training dataset (30%), in which Cox models using selected variables were fitted; and iii) hold-out test dataset (30%), in which the predictive accuracy of these models was evaluated and compared with pooled cohort equations.

### Cardiovascular disease definition

Cardiovascular disease was defined as myocardial infarction and its sequelae, angina, non-hemorrhagic stroke and transient ischemic attack. (9) Cases were identified using nurse-administered questionnaire at baseline and through linkage to hospital admissions, operation/procedure codes and death registrations (Appendix Table 1).

### Study variables

We considered variables included in PCE (3,17) (age, ethnicity [White, Black, Other], smoking [never, former, current], diabetes [prevalent self-reported or from hospital records], total and high-density lipoprotein cholesterol, systolic blood pressure [mean of two measurements] and use of antihypertensive medication), as well as additional QRISK3 variables (5) (standard deviation of systolic blood pressure, body mass index, family history of CAD, area-level deprivation score [Townsend], medication use including oral steroids and atypical antipsychotics, and self-reported prevalent conditions including chronic kidney disease stage 3–5, atrial fibrillation, migraine, rheumatoid arthritis, systemic lupus erythematosus, severe mental illness, and erectile dysfunction in men) and PRS for CVD, as previously described (9). We considered 26 other baseline serum biochemistry measurements (excluding estradiol and rheumatoid factor that were missing in more than 80% of participants) (18,19) and 23 baseline hematology measurements including full blood count and white blood cell differential (20), which were log-transformed prior to analyses due to skewed distribution (see Appendix, Methods). For biochemical and hematological variables, there was up to 20% missingness with similar proportions for CVD cases and non-cases (Appendix Table 2). Missing values were imputed using multiple imputation with predictive mean matching over five iterations of chained random forests (21).

NMR-derived metabolic variables were also available in ∼120,000 randomly sampled participants from the whole UK Biobank cohort. The high-throughput NMR profiling platform was developed by Nightingale Health Ltd. The NMR-derived metabolomic profile includes blood levels of (N=168) annotated molecules including lipoprotein lipids, fatty acids and fatty acid compositions, as well as some low-molecular weight metabolites including amino acids, ketone bodies, and glycolysis metabolites. (22) After filtering for highly correlated variables and overlap with directly measured blood markers, 18 metabolomic variables were included in our analyses (Appendix Methods, Appendix Figure 1) and were available in 68,855 of the 304,356 participants included in our study (Figure 1).

### Statistical analyses

All statistical analyses were conducted in men and women separately. We used least absolute shrinkage and selection operator (LASSO) penalized regression in a stability selection framework to identify reproducible, parsimonious sets of variables that jointly contribute to CVD risk prediction and estimate their (mutually adjusted) effect sizes. We randomly split the data into three sex-stratified and non-overlapping sets: i) a variable selection dataset (40%); ii) a training dataset (30%), in which unpenalized Cox models were fit using stably selected variables and for recalibration of PCE; and iii) a hold-out test dataset (30%), in which the predictive accuracy of these Cox models was evaluated and compared with that of PCE. We constrained the ratio of CVD cases to non-cases to be equal in all three data splits (Figure 1). The Cox models used time from baseline as the underlying time variable; outcome was CVD event, with censoring for availability of hospital admission and mortality data (7^th^ April 2021). In the subset of participants with NMR data, we carried out similar analyses and compared variable selection and model performance excluding and including metabolomic data. Pooled cohort equations were recalibrated in the training set to allow for fair comparison of models in the UK Biobank population (Appendix Methods, Appendix Figure 2).

**Figure 2.**
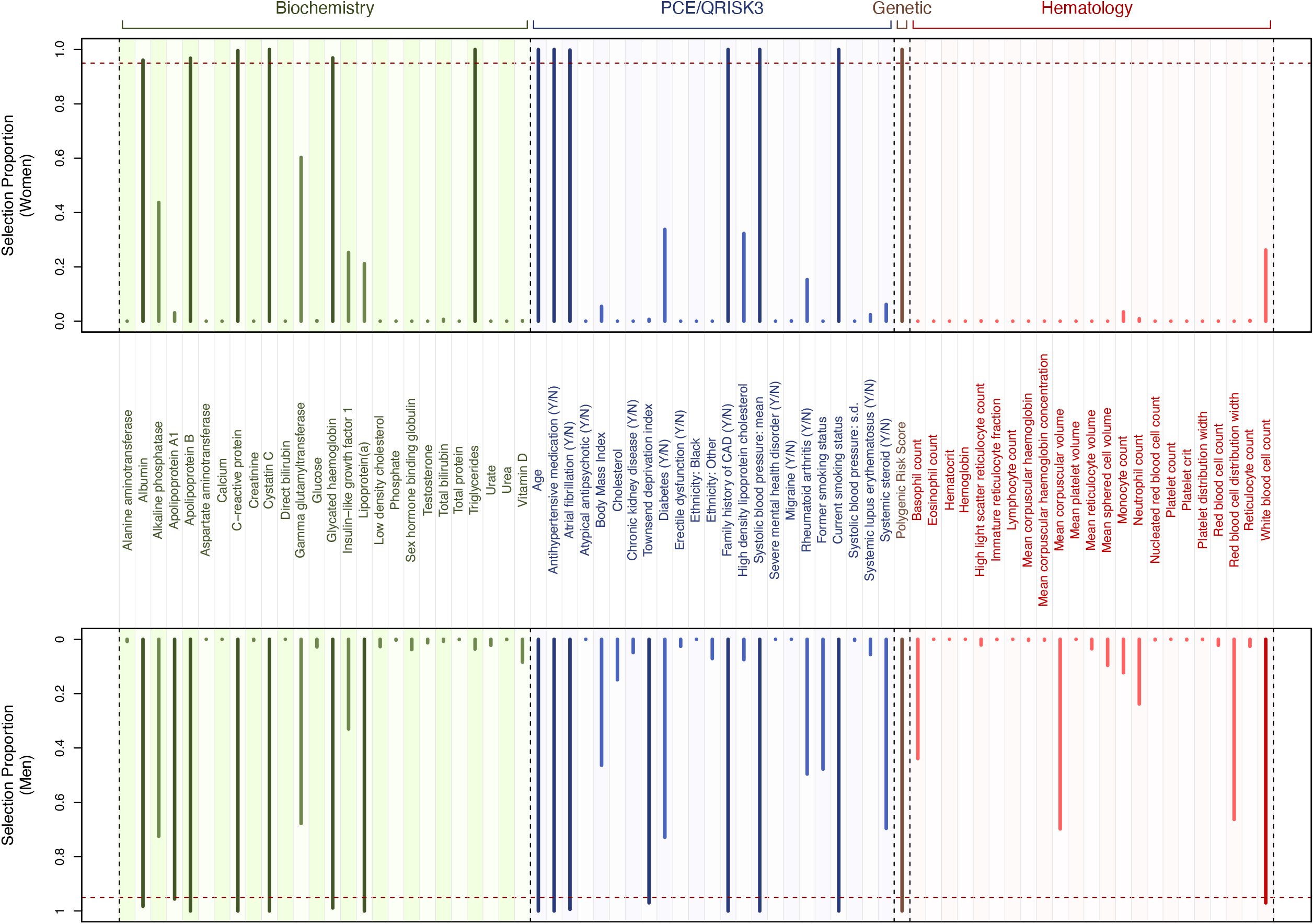
Stability selection LASSO. Selection proportions from LASSO stability selection calculated from (N=1,000) subsamples in men (A) and women (B). Explanatory variables considered include those contributing to PCE and QRISK3 scores (blue), genetic (brown), biochemical (green) and hematological (red) variables. Darker colors indicate stably selected variables (16 and 13 in men and women, respectively) as defined by variables with selection proportion above the calibrated threshold in selection proportion (vertical dark red dashed line).

### Variable selection

LASSO-regularized Cox survival models were calibrated using a stability selection approach (23). Briefly, we fit (N=1,000) LASSO Cox models on 50% subsamples of the study population and estimated, across subsamples, the per-variable selection proportion as a proxy for the variable importance. Model calibration was achieved by jointly identifying (i) the penalty parameter λ (controlling the sparsity of the LASSO model) and (ii) the threshold in selection proportion π (controlling the stability of the model, conditional on the penalty). These parameters were obtained by maximizing a likelihood-based stability score (24); features with selection proportion above the calibrated value π were considered to be stably selected.

### Predictive performance

Predictive accuracy (C-statistic) and discriminatory performance (non-nested categorical net reclassification improvement [NRI] for 7.5% 10-year risk and the associated integrated discrimination improvement [IDI], as well as the category-free NRI) (25) of Cox models with variables selected by LASSO stability selection were calculated in the test data and compared to those from recalibrated PCE. We also used a nested approach where PCE log hazards were forced into the LASSO stability selection models in place of their constituent variables.

To evaluate the contribution of each stably selected variable to model performance, we ran a series of unpenalized Cox models, sequentially adding predictors in descending order of selection proportion. These models were fit on the test set to estimate the mean and 95% confidence interval of the C-statistics.

We also used logistic models, predicting prospective CVD case status in the test set, to compare performance of regression coefficients from recalibrated PCE with LASSO stably selected variables: we performed receiver operating characteristic (ROC) analyses, reporting the mean and 95% confidence intervals of the area under the ROC curve (AUC).

In analyses of the subset of participants with NMR-derived data, we compared the predictive accuracy and discriminatory performances of models excluding and including these data.

Statistical analyses were performed using R version 4.2.2. (26)

## Results

Mean age at baseline in men was 54.4 years in non-cases and 58.9 years in cases, and 55.2 and 60.0 years respectively in women. A total of 11,899 men and 9,110 women were diagnosed with CVD during the period of follow-up (median 12.1 years). Descriptive statistics, stratified by sex and case status, are shown in Appendix Table 3, and descriptive statistics for the subset with metabolomic data in Appendix Table 4.

Our stability selection model consistently selected twelve variables in both men and women (Figure 2): age, albumin, antihypertensive medication, apolipoprotein B, atrial fibrillation, C-reactive protein, current smoker, cystatin C, family history of coronary artery disease, glycated hemoglobin, polygenic risk score (PRS) for CVD and systolic blood pressure. In addition, apolipoprotein A1, lipoprotein(a), white blood cell count and deprivation index were selected among men only, and triglycerides in women only.

Sequentially adding variables in descending order of selection proportion in unpenalized Cox models, trained and tested in separate data, showed that variables included beyond those stably selected (N=16 for men, N=13 for women) did not substantially improve model performance, as suggested by the plateauing C-statistics (Appendix Figure 3). Corresponding (mutually adjusted) regression coefficients of the unpenalized survival models are reported in Appendix Table 5.

ROC analyses with logistic models for incident CVD in test data also showed improvement in predictive accuracy when using LASSO stably selected variables versus recalibrated PCE: in men, AUCs were 0.71 [0.70-0.72] for LASSO selected variables versus 0.67 [0.66-0.68] for PCE, and in women, 0.72 [0.71-0.73] versus 0.69 [0.68-0.70], respectively (Figure 3).

**Figure 3.**
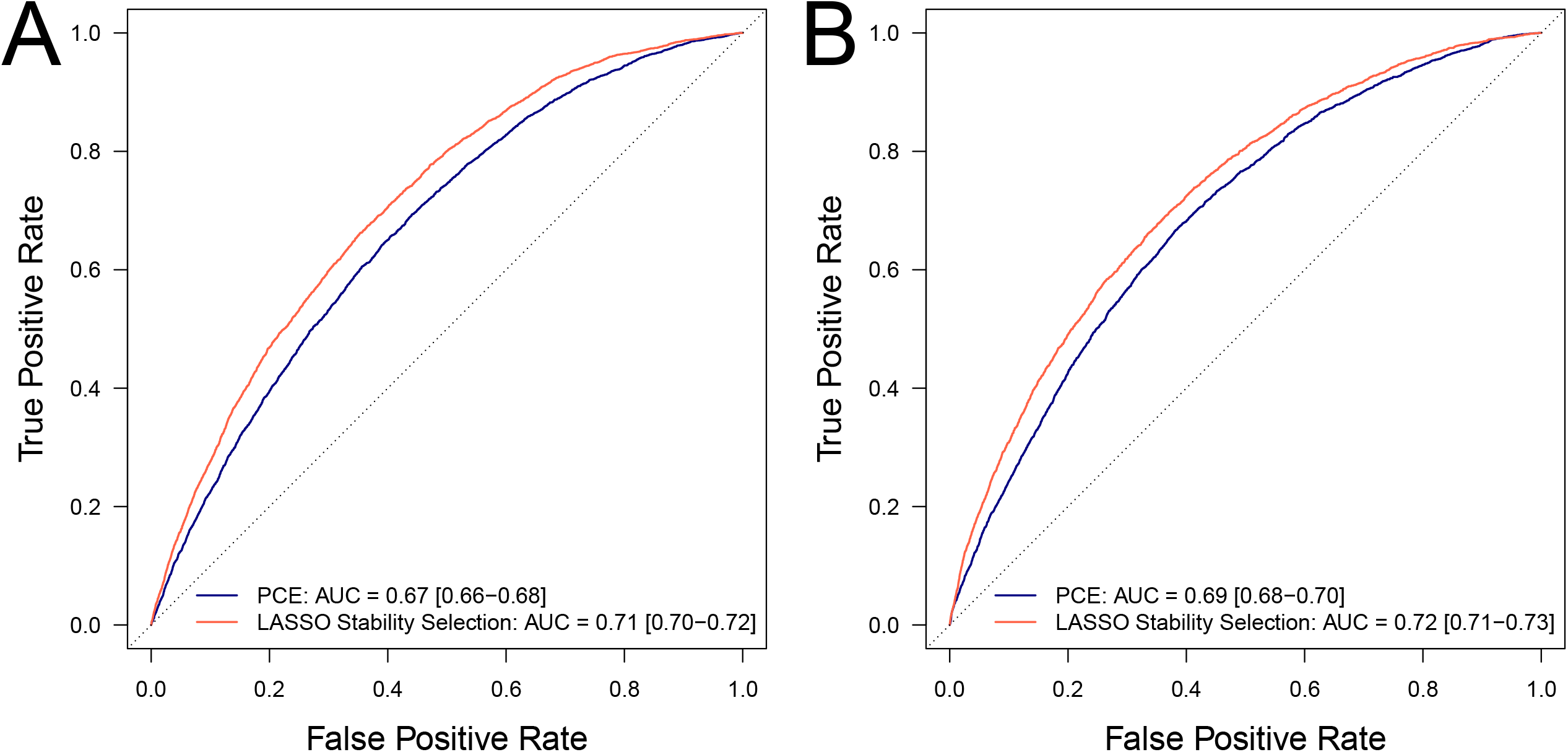
Cardiovascular disease prediction in test data. Receiver operating characteristic (ROC) curves for logistic models predicting incident CVD in men (A) and women (B) in test data, where models use either recalibrated PCE (blue line) or sex-specific LASSO stably selected variables (red line) in test data. We report the mean and 95% confidence intervals for the area under the ROC curve (AUC).

Table 1 shows the 7.5% 10-year risk prediction reclassification and categorical NRI and IDI for LASSO stability selection variables versus PCE (non-nested analyses, i.e., including individual constituents of PCE). Using LASSO stably selected variables improved classification: 35.2% of cases in men and 21.5% of cases in women were correctly reclassified as high risk (≥7.5% 10-year risk), and 27.3% of non-cases in men and 47.4% of non-cases in women were correctly reclassified as low risk (<7.5% 10-year risk). The categorical NRI and IDI were 0.054 (0.038, 0.070) and 0.018 (0.016-0.020) in men, and 0.081 (0.063, 0.099) and 0.011 (0.010-0.013) in women.

Sensitivity analyses with nested models (i.e., where PCE log hazards were included in LASSO stability selection instead of the constituent variables), stably selected slightly different variables from the main (non-nested) analyses (Appendix Figure 4), but this did not affect model performances, showing very similar C-statistics (0.70 [0.69-0.71] in men and 0.72 [0.71-0.73] in women) and similar NRI and IDI for LASSO stability selection variables alongside PCE versus PCE alone (Appendix Table 6).

Among the subset of (N=68,855) participants with available metabolomic data, LASSO stability selection identified age, antihypertensive medication, systolic blood pressure, PRS, C-reactive protein and cystatin C as jointly predictive of CVD in both men and women when metabolomic data were excluded (base models); additionally, glycated hemoglobin was selected in men only (Figure 4A) and family history of CAD and current smoking in women only (Figure 4B). Family history of CAD was also selected in men when metabolomic data were included, alongside the same variables as in the base model. In women, the model including metabolomic data also selected the same variables as in the base model except for C-reactive protein, which was substituted by glycoprotein acetyls and lipoprotein(a). There was similar predictive accuracy between stability selection models excluding and including metabolomic data, with C-statistics of 0.69 [0.67-0.71] and 0.69 [0.67-0.72] in men, and 0.73 [0.71-0.75] and 0.71 [0.71-0.75] in women, respectively (Appendix Figure 5). Logistic models for incident CVD excluding or including metabolomic data yielded similar AUCs (Appendix Figure 6).

**Figure 4.**
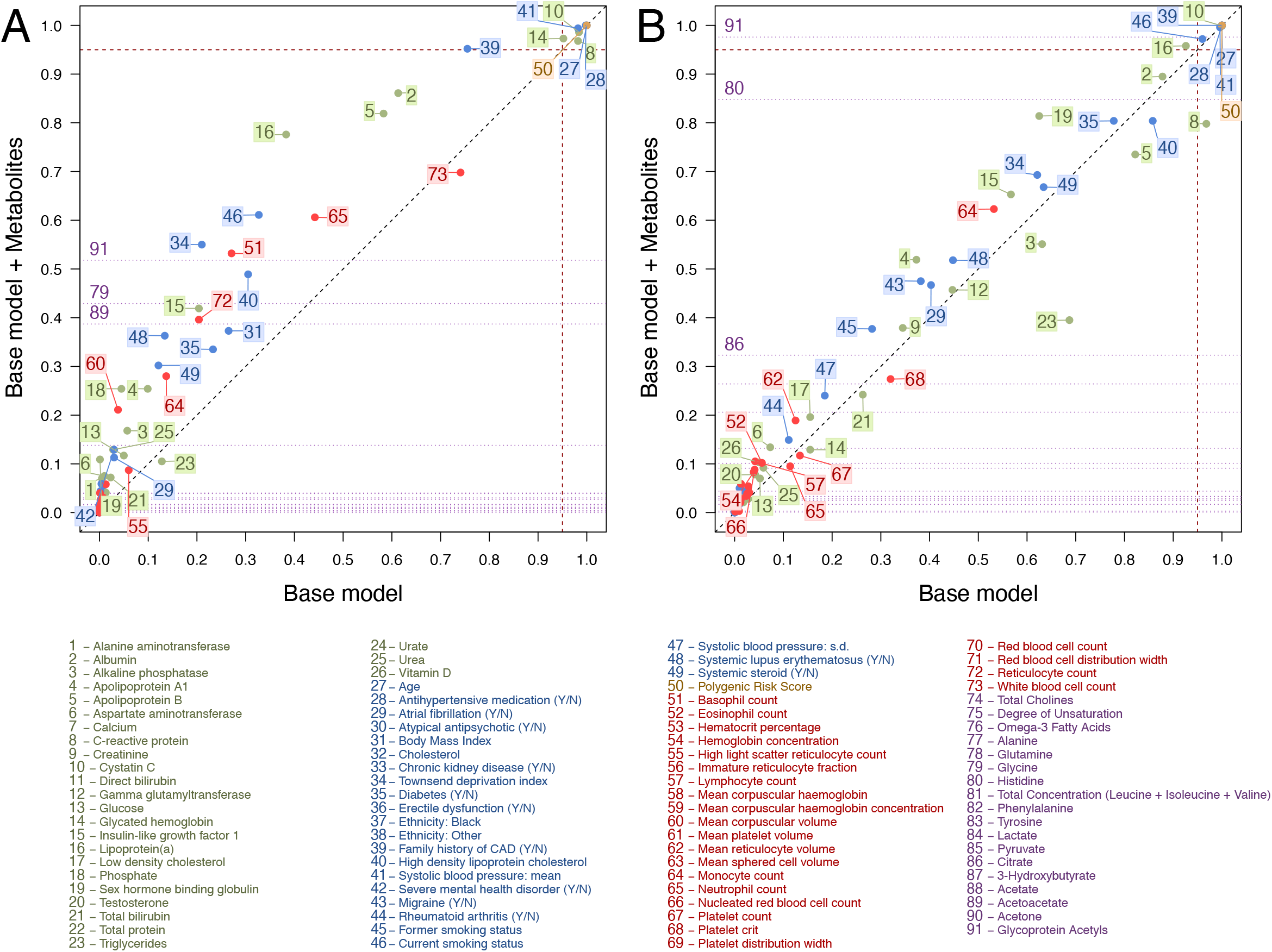
Comparison of models excluding and including metabolomic data. LASSO stability selection proportions from the model excluding (X-axis) and including (Y-axis) metabolomic data in men (A) and women (B). Calibrated thresholds in selection proportion are shown as dashed dark-red lines. In (A) and (B), variables to the right of the dashed red vertical line were selected in the base model, and variables above the horizontal dashed red line were selected in the model including metabolomic data. For display purposes, the horizontal dotted purple lines indicate the selection proportions for the metabolomic variables – only the top three metabolomic variables are numbered in each plot.

## Discussion

In this large population-based cohort, considering a wide range of demographic, lifestyle, personal/family medical history, blood-based markers including a PRS and metabolomic data, we identified parsimonious sets of variables that improved CVD risk prediction and classification beyond PCE. Inclusion of such variables could lead to better targeting of treatment for CVD prevention in the population.

A key point in interpreting our results is that variables were only stably selected if they jointly (and complementarily) predicted incident CVD. Among the variables selected for both men and women, some are already included in PCE (age, anti-hypertensive medication, smoking status and systolic blood pressure), while diabetes status, ethnicity, high-density lipoprotein and total cholesterol are included in PCE but were not selected.

Glycated hemoglobin, which captures blood glucose levels over the past two to three months (27), was selected in preference to recorded diabetes status in both men and women. Similarly, among lipids, apolipoprotein B was selected for both men and women in place of the more standard lipid measures. In addition, triglycerides (28) were selected for women only and lipoprotein(a) (29,30) and apolipoprotein A1 (31–33) for men only. PCE and QRISK3 already both include sex-specific coefficients, to account for differences in CVD risk prediction between men and women.

Apolipoprotein B has been suggested as a better predictor of CVD than standard lipid measures (34–38), but has not yet been formally incorporated into clinical risk calculators. This was however recommended by the European Society of Cardiology for risk assessment (39). In keeping, the 2019 American College of Cardiology / American Heart Association guideline on primary prevention of CVD has recommended that, if measured, apolipoprotein B level can be used to improve risk stratification among those at increased risk. (40)

Other variables selected in both men and women were family history of CAD, atrial fibrillation, C-reactive protein, albumin, cystatin C, and the PRS. Two of these variables (history of atrial fibrillation and family history of CAD) can be readily obtained from routine data linkage or questionnaire. Of the blood-based markers, three capture information about systemic inflammation and are implicated in CVD risk: serum albumin (41), C-reactive protein (42) and cystatin C (43). Both albumin and C-reactive protein are acute phase reactants whereas cystatin C is a marker of renal function (44).

The PRS was selected in all models alongside family history of CAD, suggesting that they jointly contribute to CVD risk. Family history of CAD may reflect common lifestyle and socio-economic factors as well as genetic risk (29). Although PRS was selected for both men and women, it made only modest contribution to predictive accuracy, in keeping with previous analyses of UK Biobank and other data.(9,45,46)

NMR-derived metabolomic data did not improve predictive accuracy above the other variables, and in women only glycoprotein acetyls (47–49) was selected in preference to C-reactive protein, which did not affect predictive accuracy. These two variables are highly correlated and while C-reactive protein is routinely measured, this is currently not the case for NMR-derived measures. No metabolomic variables were stably selected for men.

### Limitations

Our study has limitations. First, it only included participants aged 38 to 73 years at baseline who were mostly of European ancestry; the participants were on average healthier, less deprived and have lower mortality than the general population and therefore may not be fully representative. (50) Second, the variable selection, training and test data were drawn from the same population; external validation in different cohorts and settings is needed to generalize our findings to other populations. Third, while PCE were developed in US cohorts, the present study uses a UK-based cohort; we performed model recalibration to correct for population differences. (51) Fourth, other potentially important predictors including coronary artery calcium were not available in our data and may further improve risk prediction or potentially compete with variables our models have selected (52). Finally, cost-benefit and decision analyses would be needed before an enhanced predictive score was widely adopted into clinical practice.

## Conclusions

While addition of single biomarkers or PRS has shown modest improvements in risk accuracy, our results show greater improvements when multiple self-reported and other readily available data are considered in combination. The selected variables should be considered in future re-evaluation of risk assessment strategies for CVD.

## Data Availability

This study was conducted using the UK Biobank resource under application number 69328 granting access to the corresponding UK Biobank genetic and phenotype data.

## Appendix material

Methods

Appendix Tables 1-6

Appendix Figures 1-6

## Data Availability

This study was conducted using the UK Biobank resource under application number 69328 granting access to the corresponding UK Biobank genetic and phenotype data. The UK Biobank received ethical approval from the North West Multi-centre Research Ethics Committee (REC reference: 11/NW/0382) to obtain and disseminate data and samples from the participants (http://www.ukbiobank.ac.uk/ethics/).

## Acknowledgements

We thank Dr Verena Zuber for helpful discussions.

## Declaration of Helsinki

The study complies with the Declaration of Helsinki.

