## Supplementary methods, figures and tables for "Improving cardiovascular risk prediction beyond pooled cohort equations: a prospective cohort of 304,356 participants"

**APPENDIX**

Elliott J, Bodinier B, Whitaker M, et al. Cardiovascular risk prediction using variable selection and ranking in a prospective cohort of 304,356 participants

**Methods**

*Variable definitions*

Self-reported ethnicity was defined from UK Biobank field 21000 to reproduce ethnicity categories used in pooled cohort equations (White, Black, Other). Pooled cohort equations compute cardiovascular risk using the same weightings for White and Other ethnicity as well as for individuals not entering their ethnicity, with Black ethnicity having a different set of weightings. Black ethnicity included codes 4, 2001, 2002, 4001, 4002 and 4003. White ethnicity included codes 1, 1001, 1002 and 1003. If UK Biobank data field 21000 had missing data, was answered "Do not know" or "Prefer not to answer", ethnicity was coded as Other.

Diabetes in pooled cohort equations is a binary variable (Y/N). Here, "yes" included any type of diabetes mellitus according to:

- self-report (UK Biobank field 20002 – codes: 1220, 1222, 1223)
- self-reported diabetic medication use (UK Biobank field 20003 – codes: 1140868902, 1140874646, 1140874674, 1140874718, 1140874744, 1140883066, 1140884600, 1141152590, 1141157284, 1141168660, 1141171646, 1141173882, 1141189090)
- hospitalization prior to enrolment (ICD-9 codes: 25000, 25010, 25020, 25030, 25040, 25050, 25060, 25070, 25080, 25090, 25002, 25012, 25022, 25032, 25042, 25052, 25062, 25072, 25082, 25092, 25001, 25011, 25021, 25031, 25041, 25051, 25061, 25071, 25081, 25091, 25003, 25013, 25023, 25033, 25043, 25053, 25063, 25073, 25083, 25093. ICD-10 codes: E10, E11, O230, O231.
- glycated hemoglobin at baseline was not used to define diabetes to avoid capturing unknown cases.

Smoking status (never, former, current) was defined using UK Biobank fields 20116, 22508, 3456, 1239, 22506, 22508 and 3446.

*Variable filtering*

Circulating levels of 168 metabolomic biomarkers, including fatty acids, cholesterol, lipoproteins, phospholipids, amino acids and ketone bodies, were measured in a subset of N=117,413 UK Biobank participants at baseline (N=73,345 in our study population). For the N=5 biomarkers (albumin, apolipoprotein A1, apolipoprotein B, creatinine and glucose) measured on both the Nightingale and biochemistry platforms, only biochemistry measurements were retained for analysis. All lipid-related markers (N=125) were excluded due to strong correlations with each other or with biochemistry variables. Due to strong correlations ($\rho$>0.7) with biochemistry biomarkers, we excluded an additional set of 18 metabolomic markers. The total concentration of branched-chain amino acids was used instead of its individual components (Valine, Leucine and Isoleucine) which were strongly correlated ($\rho$>0.8). Similarly, phosphatidylcholines and phosphoglycerides, which were both strongly correlated with total cholines ($\rho$>0.9) were excluded. After filtering, a total of 18 metabolomic features, with low to moderate correlations with each other and with biochemistry and hematology markers, remained for analysis (Appendix Figure 1). Complete data for the 18 metabolomic markers were available for N=68,855 participants.

*Recalibration of PCE*

Pooled cohort equations were recalibrated in the training set in men and women separately to correct for possible over-estimation of CVD risk in UK Biobank and to allow for fair comparison between models. Recalibration was achieved by fitting predicted log hazards from published PCE as a Cox model covariate, with baseline survival function estimated to give the intercept. Calibration was assessed graphically by plotting observed probability of CVD (Kaplan-Meier estimate) against mean predicted probability within decile of predicted probabilities. We calculated the calibration slope to assess possible differences between observed and expected probabilities (Appendix Figure 2).

**Appendix Table 1.** Case definition for cardiovascular disease

| ICD-10 | ICD-9 | OPCS-4 | Biobank field: 20002 | Biobank field: 20004 | Biobank field: 6150 |
|---|---|---|---|---|---|
| G45 | 410 | K40 | 1074: Angina | 1070: Coronary angioplasty | 1: Heart attack |
| I20 | 411 | K41 | 1075: Heart attack/myocardial infarction | 1071: Other arterial surgery/revascularization procedures | 2: Angina |
| I21 | 412 | K42 | 1082: Transient ischemic attack | 1095: Coronary artery bypass grafts | 3: Stroke |
| I22 | 413 | K43 | 1583: Ischemic stroke | 1105: Carotid artery surgery/endarterectomy | |
| I23 | 414 | K44 | | 1109: Carotid artery angioplasty +/- stent | |
| I24 | 434 | K45 | | 1514: Coronary angiogram | |
| I25 | 436 | K46 | | | |
| I63 | | K47.1 | | | |
| I64 | | K49 | | | |
| | | K50 | | | |
| | | K75 | | | |

**Appendix Table 2.** Missingness (%) by variable prior to imputation, stratified by case status and sex.

| | *Men* | | *Women* | |
|---|---|---|---|---|
| **Variable** | **Cases** | **Non-cases** | **Cases** | **Non-cases** |
| Alanine aminotransferase (U/L) | 0.09 | 0.09 | 0.01 | 0.03 |
| Albumin (g/L) | 0.04 | 0.04 | 0.04 | 0.04 |
| Alkaline phosphatase (U/L) | 0.02 | 0.02 | 0.01 | 0.01 |
| Apolipoprotein A1 (g/L) | 0.29 | 0.18 | 1.00 | 0.94 |
| Apolipoprotein B (g/L) | 0.73 | 0.66 | 0.53 | 0.36 |
| Aspartate aminotransferase (U/L) | 0.45 | 0.41 | 0.41 | 0.40 |
| Basophil count (e9 cells/L) | 2.40 | 2.49 | 2.93 | 2.81 |
| C-reactive protein (mg/L) | 0.33 | 0.25 | 0.26 | 0.18 |
| Calcium (mmol/L) | 0.07 | 0.07 | 0.08 | 0.07 |
| Creatinine (µmol/L) | 0.10 | 0.07 | 0.07 | 0.07 |
| Cystatin C (mg/L) | 0.06 | 0.07 | 0.05 | 0.05 |
| Direct bilirubin (µmol/L) | 8.57 | 7.21 | 27.67 | 22.26 |
| Eosinophil count (e9 cells/L) | 2.40 | 2.49 | 2.93 | 2.81 |
| Gamma glutamyltransferase (U/L) | 0.12 | 0.07 | 0.07 | 0.06 |
| Glucose (mmol/L) | 0.12 | 0.13 | 0.08 | 0.11 |
| Glycated hemoglobin (mmol/mol) | 4.48 | 4.85 | 4.96 | 4.78 |
| Hematocrit (%) | 2.27 | 2.33 | 2.78 | 2.63 |
| Hemoglobin concentration (g/dL) | 2.27 | 2.33 | 2.78 | 2.63 |
| High light scatter reticulocyte count (e12 cells/L) | 3.46 | 3.88 | 4.22 | 4.28 |
| Immature reticulocyte fraction | 3.46 | 3.88 | 4.22 | 4.28 |
| Insulin-like growth factor 1 (nmol/L) | 0.57 | 0.59 | 0.83 | 0.63 |
| Lipoprotein(a) (g/L) | 19.50 | 19.19 | 20.02 | 18.98 |
| Low-density lipoprotein cholesterol (mmol/L) | 0.18 | 0.17 | 0.15 | 0.16 |
| Lymphocyte count (e9 cells/L) | 2.40 | 2.49 | 2.93 | 2.81 |
| Mean corpuscular hemoglobin (pg) | 2.27 | 2.33 | 2.78 | 2.63 |
| Mean corpuscular hemoglobin concentration (g/dL) | 2.27 | 2.33 | 2.78 | 2.63 |
| Mean corpuscular volume (fL) | 2.27 | 2.33 | 2.78 | 2.63 |
| Mean platelet volume (fL) | 2.27 | 2.33 | 2.78 | 2.63 |
| Mean reticulocyte volume (fL) | 3.46 | 3.88 | 4.22 | 4.28 |
| Mean sphered cell volume (fL) | 3.46 | 3.88 | 4.22 | 4.28 |
| Monocyte count (e9 cells/L) | 2.40 | 2.49 | 2.93 | 2.81 |
| Neutrophil count (e9 cells/L) | 2.40 | 2.49 | 2.93 | 2.81 |
| Nucleated red blood cell count (e9 cells/L) | 2.40 | 2.50 | 2.93 | 2.81 |
| Phosphate (mmol/L) | 0.29 | 0.23 | 0.25 | 0.23 |
| Platelet count (e9 cells/L) | 2.27 | 2.33 | 2.78 | 2.63 |
| Platelet crit (%) | 2.27 | 2.33 | 2.78 | 2.63 |
| Platelet distribution width (%) | 2.27 | 2.33 | 2.78 | 2.63 |
| Red blood cell count (e12 cells/L) | 2.27 | 2.33 | 2.78 | 2.63 |
| Red blood cell distribution width (%) | 2.27 | 2.33 | 2.78 | 2.63 |
| Reticulocyte count (e9 cells/L) | 3.46 | 3.88 | 4.22 | 4.28 |
| Sex hormone binding globulin (nmol/L) | 0.69 | 0.86 | 1.20 | 1.23 |
| Testosterone (nmol/L) | 1.03 | 0.94 | 19.78 | 15.09 |
| Total bilirubin (µmol/L) | 0.50 | 0.43 | 0.44 | 0.44 |
| Total protein (g/L) | 0.18 | 0.16 | 0.15 | 0.13 |
| Triglycerides (mmol/L) | 0.11 | 0.08 | 0.04 | 0.07 |

| | | | | |
|---|---|---|---|---|
| Urate (μmol/L) | 0.13 | 0.11 | 0.10 | 0.13 |
| Urea (mmol/L) | 0.13 | 0.09 | 0.07 | 0.09 |
| Vitamin D (μg) | 5.19 | 3.64 | 9.69 | 6.20 |
| White blood cell count (e9 cells/L) | 2.27 | 2.33 | 2.78 | 2.63 |

**Appendix Table 3.** Descriptive statistics of data with imputation, stratified by case status for (A) men and (B) women. For continuous variables, mean values (s.d.) shown and for binary variables, prevalence of “yes” (%) is shown.

**(A)**

| **Variable** | **Non-cases** | **Cases** |
|---|---|---|
| *N* | 109,825 | 11,899 |
| | **Mean (s.d.)** | |
| Age (years) | 54.40 (8.30) | 58.88 (7.40) |
| Body mass index (kg/m2) | 27.30 (3.98) | 27.98 (4.20) |
| Systolic blood pressure: mean (mmHg) | 139.58 (17.04) | 145.78 (18.30) |
| Systolic blood pressure: s.d. (mmHg) | 5.13 (4.25) | 5.49 (4.50) |
| Townsend deprivation index | -1.37 (3.07) | -1.23 (3.17) |
| Alanine aminotransferase (U/L) | 27.14 (15.48) | 26.84 (15.64) |
| Albumin (g/L) | 45.65 (2.57) | 45.05 (2.59) |
| Alkaline phosphatase (U/L) | 81.18 (23.58) | 84.88 (28.54) |
| Apolipoprotein A1 (g/L) | 1.44 (0.23) | 1.41 (0.23) |
| Apolipoprotein B (g/L) | 1.07 (0.23) | 1.12 (0.23) |
| Aspartate aminotransferase (U/L) | 27.93 (11.42) | 28.08 (11.77) |
| Calcium (mmol/L) | 2.37 (0.09) | 2.37 (0.09) |
| Cholesterol (mmol/L) | 222.27 (38.91) | 228.67 (40.18) |
| C-reactive protein (mg/L) | 2.31 (4.13) | 3.07 (4.91) |
| Creatinine (µmol/L) | 80.70 (13.56) | 81.75 (20.84) |
| Cystatin C (mg/L) | 0.92 (0.14) | 0.97 (0.18) |
| Direct bilirubin (µmol/L) | 1.93 (0.89) | 1.88 (0.90) |
| Gamma glutamyltransferase (U/L) | 43.25 (45.33) | 47.41 (50.93) |
| Glucose (mmol/L) | 5.00 (0.99) | 5.10 (1.25) |
| Glycated hemoglobin (mmol/mol) | 34.95 (5.38) | 36.23 (6.71) |
| High-density lipoprotein cholesterol (mmol/L) | 50.41 (11.97) | 49.00 (11.94) |
| Insulin-like growth factor 1 (nmol/L) | 22.33 (5.41) | 21.48 (5.50) |
| Lipoprotein(a) (g/L) | 42.36 (47.75) | 46.76 (49.19) |
| Low density cholesterol (mmol/L) | 3.70 (0.77) | 3.84 (0.80) |
| Phosphate (mmol/L) | 1.11 (0.16) | 1.11 (0.16) |
| Sex hormone binding globulin (nmol/L) | 39.43 (16.62) | 41.57 (17.56) |
| Testosterone (nmol/L) | 12.34 (3.70) | 12.18 (3.78) |
| Total bilirubin (µmol/L) | 10.34 (4.90) | 10.03 (4.60) |
| Total protein (g/L) | 72.66 (4.04) | 72.50 (4.16) |
| Triglycerides (mmol/L) | 1.94 (1.14) | 2.08 (1.19) |
| Urate (µmol/L) | 350.83 (68.56) | 358.73 (72.64) |
| Urea (mmol/L) | 5.48 (1.26) | 5.63 (1.48) |
| Vitamin D (µg) | 48.04 (20.99) | 47.60 (20.68) |
| Basophil count (e9 cells/L) | 0.03 (0.04) | 0.04 (0.05) |
| Eosinophil count (e9 cells/L) | 0.18 (0.14) | 0.19 (0.15) |
| Hematocrit (%) | 43.47 (2.87) | 43.61 (3.08) |
| Hemoglobin concentration (g/dL) | 15.06 (0.97) | 15.10 (1.04) |
| High light scatter reticulocyte count (e12 cells/L) | 0.02 (0.01) | 0.02 (0.01) |
| Immature reticulocyte fraction | 0.28 (0.06) | 0.29 (0.06) |
| Lymphocyte count (e9 cells/L) | 1.88 (1.29) | 1.96 (2.06) |
| Mean corpuscular hemoglobin (pg) | 31.64 (1.79) | 31.78 (1.83) |
| Mean corpuscular hemoglobin concentration (g/dL) | 34.67 (1.03) | 34.64 (1.03) |

| | | |
|---|---|---|
| Mean corpuscular volume (fL) | 91.28 (4.37) | 91.75 (4.54) |
| Mean platelet volume (fL) | 9.27 (1.06) | 9.25 (1.06) |
| Mean reticulocyte volume (fL) | 106.09 (7.65) | 106.84 (7.82) |
| Mean sphered cell volume (fL) | 82.72 (5.26) | 83.23 (5.44) |
| Monocyte count (e9 cells/L) | 0.50 (0.21) | 0.53 (0.25) |
| Neutrophil count (e9 cells/L) | 4.13 (1.39) | 4.40 (1.47) |
| Nucleated red blood cell count (e9 cells/L) | 0.02 (0.03) | 0.02 (0.02) |
| Platelet count (e9 cells/L) | 239.43 (54.72) | 239.64 (58.49) |
| Platelet crit (%) | 0.22 (0.04) | 0.22 (0.05) |
| Platelet distribution width (%) | 16.55 (0.52) | 16.58 (0.53) |
| Red blood cell count (e12 cells/L) | 4.77 (0.37) | 4.76 (0.39) |
| Red blood cell distribution width (%) | 13.39 (0.83) | 13.52 (0.89) |
| Reticulocyte count (e9 cells/L) | 0.06 (0.04) | 0.07 (0.04) |
| White blood cell count (e9 cells/L) | 6.73 (2.13) | 7.12 (2.76) |
| Polygenic risk score | -0.03 (0.78) | 0.15 (0.79) |
| | **Prevalence of "yes" (%)** | |
| Antihypertensive medication | 12,546 (11.42) | 2,501 (21.02) |
| Atrial fibrillation | 888 (0.81) | 250 (2.10) |
| Atypical antipsychotic | 290 (0.26) | 29 (0.24) |
| Chronic kidney disease (stages 3-5) | 45 (0.04) | 15 (0.13) |
| Diabetes | 2,347 (2.14) | 540 (4.54) |
| Erectile dysfunction | 463 (0.42) | 89 (0.75) |
| Ethnicity: Black | 2,014 (1.83) | 119 (1.00) |
| Ethnicity: Other | 4,251 (3.87) | 428 (3.60) |
| Family history of coronary artery disease | 38,491 (35.05) | 5,241 (44.05) |
| Migraine | 1,733 (1.58) | 179 (1.50) |
| Rheumatoid arthritis | 627 (0.57) | 148 (1.24) |
| Severe mental health disorder | 503 (0.46) | 61 (0.51) |
| Smoker: current | 12,473 (11.36) | 1,809 (15.20) |
| Smoker: former | 38,870 (35.39) | 4,857 (40.82) |
| Systemic lupus erythematosus | 29 (0.03) | 8 (0.07) |
| Systemic steroid | 690 (0.63) | 149 (1.25) |

**(B)**

| Variable | Non-cases | Cases |
|---|---|---|
| *N* | 173,522 | 9,110 |
| | Mean (s.d.) | |
| Age (years) | 55.15 (7.98) | 60.00 (6.89) |
| Body mass index (kg/m2) | 26.59 (4.92) | 27.86 (5.33) |
| Systolic blood pressure: mean (mmHg) | 133.85 (18.96) | 142.22 (20.12) |
| Systolic blood pressure: s.d. (mmHg) | 5.40 (4.45) | 5.86 (4.84) |
| Townsend deprivation index | -1.46 (2.97) | -1.22 (3.10) |
| Alanine aminotransferase (U/L) | 19.52 (11.86) | 20.97 (12.62) |
| Albumin (g/L) | 44.98 (2.57) | 44.51 (2.63) |
| Alkaline phosphatase (U/L) | 83.07 (26.76) | 91.26 (29.32) |
| Apolipoprotein A1 (g/L) | 1.65 (0.27) | 1.61 (0.28) |
| Apolipoprotein B (g/L) | 1.05 (0.23) | 1.13 (0.24) |
| Aspartate aminotransferase (U/L) | 24.04 (9.30) | 25.15 (10.22) |
| Calcium (mmol/L) | 2.38 (0.10) | 2.39 (0.10) |
| Cholesterol (mmol/L) | 231.02 (41.72) | 242.86 (42.61) |
| C-reactive protein (mg/L) | 2.56 (4.17) | 3.55 (5.07) |
| Creatinine (μmol/L) | 63.77 (10.76) | 64.81 (18.65) |
| Cystatin C (mg/L) | 0.86 (0.14) | 0.93 (0.20) |
| Direct bilirubin (μmol/L) | 1.53 (0.65) | 1.45 (0.62) |
| Gamma glutamyltransferase (U/L) | 28.65 (31.78) | 34.49 (38.78) |
| Glucose (mmol/L) | 4.97 (0.81) | 5.11 (1.10) |
| Glycated hemoglobin (mmol/mol) | 34.94 (4.56) | 36.49 (5.89) |
| High-density lipoprotein cholesterol (mmol/L) | 62.44 (14.45) | 59.71 (14.39) |
| Insulin-like growth factor 1 (nmol/L) | 21.27 (5.72) | 19.78 (5.68) |
| Lipoprotein(a) (g/L) | 44.74 (49.02) | 48.22 (50.32) |
| Low density cholesterol (mmol/L) | 3.71 (0.84) | 3.98 (0.85) |
| Phosphate (mmol/L) | 1.19 (0.15) | 1.20 (0.15) |
| Sex hormone binding globulin (nmol/L) | 64.41 (31.45) | 60.76 (30.85) |
| Testosterone (nmol/L) | 1.11 (0.62) | 1.14 (0.67) |
| Total bilirubin (μmol/L) | 8.16 (3.70) | 7.80 (3.41) |
| Total protein (g/L) | 72.40 (4.09) | 72.33 (4.22) |
| Triglycerides (mmol/L) | 1.49 (0.82) | 1.78 (0.94) |
| Urate (μmol/L) | 264.68 (62.27) | 282.77 (68.47) |
| Urea (mmol/L) | 5.13 (1.24) | 5.42 (1.41) |
| Vitamin D (μg) | 48.60 (20.79) | 47.10 (20.29) |
| Basophil count (e9 cells/L) | 0.04 (0.05) | 0.04 (0.06) |
| Eosinophil count (e9 cells/L) | 0.16 (0.13) | 0.17 (0.13) |
| Hematocrit (%) | 39.16 (2.79) | 39.55 (2.88) |
| Hemoglobin concentration (g/dL) | 13.47 (0.96) | 13.60 (0.98) |
| High light scatter reticulocyte count (e12 cells/L) | 0.02 (0.01) | 0.02 (0.01) |
| Immature reticulocyte fraction | 0.29 (0.06) | 0.30 (0.06) |
| Lymphocyte count (e9 cells/L) | 1.99 (0.94) | 2.08 (0.76) |
| Mean corpuscular hemoglobin (pg) | 31.29 (1.95) | 31.32 (1.96) |
| Mean corpuscular hemoglobin concentration (g/dL) | 34.41 (1.05) | 34.41 (1.10) |
| Mean corpuscular volume (fL) | 90.90 (4.69) | 90.99 (4.69) |
| Mean platelet volume (fL) | 9.37 (1.09) | 9.33 (1.09) |
| Mean reticulocyte volume (fL) | 105.57 (7.75) | 105.94 (8.08) |
| Mean sphered cell volume (fL) | 83.10 (5.30) | 83.24 (5.55) |
| Monocyte count (e9 cells/L) | 0.43 (0.20) | 0.46 (0.20) |

| | | |
|---|---|---|
| Neutrophil count (e9 cells/L) | 4.14 (1.37) | 4.37 (1.51) |
| Nucleated red blood cell count (e9 cells/L) | 0.02 (0.04) | 0.02 (0.02) |
| Platelet count (e9 cells/L) | 266.18 (60.12) | 270.53 (63.78) |
| Platelet crit (%) | 0.25 (0.05) | 0.25 (0.05) |
| Platelet distribution width (%) | 16.42 (0.51) | 16.44 (0.51) |
| Red blood cell count (e12 cells/L) | 4.31 (0.33) | 4.35 (0.35) |
| Red blood cell distribution width (%) | 13.51 (1.06) | 13.61 (1.08) |
| Reticulocyte count (e9 cells/L) | 0.06 (0.03) | 0.06 (0.04) |
| White blood cell count (e9 cells/L) | 6.76 (1.88) | 7.10 (1.91) |
| Polygenic risk score | 0.00 (0.79) | 0.15 (0.80) |
| | **Prevalence of "yes" (%)** | |
| Antihypertensive medication | 20,247 (11.67) | 2,221 (24.38) |
| Atrial fibrillation | 561 (0.32) | 140 (1.54) |
| Atypical antipsychotic | 339 (0.20) | 26 (0.29) |
| Chronic kidney disease (stages 3-5) | 47 (0.03) | 11 (0.12) |
| Diabetes | 2,165 (1.25) | 281 (3.08) |
| Ethnicity: Black | 3,205 (1.85) | 123 (1.35) |
| Ethnicity: Other | 5,947 (3.43) | 302 (3.32) |
| Family history of CAD | 73,224 (42.20) | 4,895 (53.73) |
| Migraine | 7,701 (4.44) | 446 (4.90) |
| Rheumatoid arthritis | 2,227 (1.28) | 233 (2.56) |
| Severe mental health disorder | 694 (0.40) | 64 (0.70) |
| Smoker: current | 13,447 (7.75) | 1,146 (12.58) |
| Smoker: former | 54,916 (31.65) | 3,160 (34.69) |
| Systemic lupus erythematosus | 314 (0.18) | 45 (0.49) |
| Systemic steroid | 1,306 (0.75) | 156 (1.71) |

**Appendix Table 4.** Descriptive statistics for the subset with NMR-derived metabolomic data, stratified by case status, for (A) men and (B) women. For continuous variables, mean values (s.d.) shown and for binary variables, prevalence of "yes" (%) is shown.

**(A)**

| **Variable** | **Non-cases** | **Cases** |
|---|---|---|
| *N* | 25,205 | 2,668 |
| | **Mean (s.d.)** | |
| Age (years) | 54.36 (8.30) | 58.92 (7.41) |
| Body mass index (kg/m2) | 27.30 (3.97) | 27.99 (4.28) |
| Systolic blood pressure: mean (mmHg) | 139.54 (16.96) | 145.73 (18.40) |
| Systolic blood pressure: s.d. (mmHg) | 5.14 (4.23) | 5.52 (4.56) |
| Townsend deprivation index | -1.40 (3.07) | -1.25 (3.15) |
| Alanine aminotransferase (U/L) | 27.26 (16.03) | 26.35 (15.30) |
| Albumin (g/L) | 45.69 (2.55) | 45.04 (2.59) |
| Alkaline phosphatase (U/L) | 81.02 (23.81) | 83.51 (23.89) |
| Apolipoprotein A1 (g/L) | 1.43 (0.23) | 1.41 (0.23) |
| Apolipoprotein B (g/L) | 1.07 (0.23) | 1.12 (0.23) |
| Aspartate aminotransferase (U/L) | 27.88 (12.06) | 27.63 (10.71) |
| Calcium (mmol/L) | 2.37 (0.09) | 2.37 (0.09) |
| Cholesterol (mmol/L) | 222.15 (38.96) | 227.51 (39.15) |
| C-reactive protein (mg/L) | 2.31 (4.18) | 3.00 (4.92) |
| Creatinine (µmol/L) | 80.85 (13.45) | 81.53 (14.06) |
| Cystatin C (mg/L) | 0.92 (0.13) | 0.97 (0.16) |
| Direct bilirubin (µmol/L) | 1.94 (0.93) | 1.85 (0.76) |
| Gamma glutamyltransferase (U/L) | 42.72 (41.52) | 46.00 (46.09) |
| Glucose (mmol/L) | 5.00 (0.95) | 5.11 (1.32) |
| Glycated hemoglobin (mmol/mol) | 34.87 (5.21) | 36.16 (6.64) |
| High-density lipoprotein cholesterol (mmol/L) | 50.38 (11.89) | 48.95 (11.96) |
| Insulin-like growth factor 1 (nmol/L) | 22.37 (5.37) | 21.44 (5.35) |
| Lipoprotein(a) (g/L) | 42.52 (47.88) | 47.29 (49.46) |
| Low density cholesterol (mmol/L) | 3.70 (0.77) | 3.82 (0.78) |
| Phosphate (mmol/L) | 1.11 (0.16) | 1.11 (0.16) |
| Sex hormone binding globulin (nmol/L) | 39.42 (16.53) | 41.76 (17.54) |
| Testosterone (nmol/L) | 12.36 (3.69) | 12.17 (3.83) |
| Total bilirubin (µmol/L) | 10.37 (4.96) | 9.89 (4.29) |
| Total protein (g/L) | 72.64 (4.03) | 72.44 (4.21) |
| Triglycerides (mmol/L) | 1.92 (1.12) | 2.05 (1.18) |
| Urate (µmol/L) | 350.62 (68.05) | 357.56 (73.32) |
| Urea (mmol/L) | 5.49 (1.26) | 5.65 (1.36) |
| Vitamin D (µg) | 47.92 (20.86) | 48.10 (20.60) |
| Basophil count (e9 cells/L) | 0.03 (0.04) | 0.04 (0.05) |
| Eosinophil count (e9 cells/L) | 0.18 (0.14) | 0.19 (0.15) |
| Hematocrit (%) | 43.45 (2.85) | 43.48 (3.11) |
| Hemoglobin concentration (g/dL) | 15.06 (0.97) | 15.07 (1.05) |
| High light scatter reticulocyte count (e12 cells/L) | 0.02 (0.01) | 0.02 (0.01) |
| Immature reticulocyte fraction | 0.28 (0.06) | 0.29 (0.06) |
| Lymphocyte count (e9 cells/L) | 1.88 (1.31) | 1.94 (1.33) |
| Mean corpuscular hemoglobin (pg) | 31.64 (1.77) | 31.79 (2.02) |

| | | |
|---|---|---|
| Mean corpuscular hemoglobin concentration (g/dL) | 34.68 (1.02) | 34.68 (1.14) |
| Mean corpuscular volume (fL) | 91.26 (4.35) | 91.66 (4.69) |
| Mean platelet volume (fL) | 9.27 (1.07) | 9.24 (1.06) |
| Mean reticulocyte volume (fL) | 106.11 (7.57) | 106.81 (8.00) |
| Mean sphered cell volume (fL) | 82.67 (5.16) | 83.14 (5.58) |
| Monocyte count (e9 cells/L) | 0.50 (0.21) | 0.53 (0.24) |
| Neutrophil count (e9 cells/L) | 4.12 (1.40) | 4.41 (1.46) |
| Nucleated red blood cell count (e9 cells/L) | 0.02 (0.04) | 0.02 (0.02) |
| Platelet count (e9 cells/L) | 239.72 (55.03) | 239.16 (56.65) |
| Platelet crit (%) | 0.22 (0.04) | 0.22 (0.05) |
| Platelet distribution width (%) | 16.56 (0.52) | 16.59 (0.51) |
| Red blood cell count (e12 cells/L) | 4.77 (0.37) | 4.75 (0.40) |
| Red blood cell distribution width (%) | 13.38 (0.82) | 13.51 (0.92) |
| Reticulocyte count (e9 cells/L) | 0.06 (0.03) | 0.06 (0.03) |
| White blood cell count (e9 cells/L) | 6.71 (2.11) | 7.12 (2.28) |
| 3-Hydroxybutyrate (mmol/l) | 0.06 (0.06) | 0.06 (0.06) |
| Acetate (mmol/l) | 0.02 (0.01) | 0.02 (0.01) |
| Acetoacetate (mmol/l) | 0.01 (0.01) | 0.01 (0.01) |
| Acetone (mmol/l) | 0.01 (0.01) | 0.01 (0.01) |
| Alanine (mmol/l) | 0.30 (0.07) | 0.30 (0.07) |
| Citrate (mmol/l) | 0.06 (0.01) | 0.06 (0.01) |
| Fatty acids: degree of unsaturation | 1.34 (0.08) | 1.33 (0.08) |
| Glutamine (mmol/l) | 0.54 (0.08) | 0.54 (0.08) |
| Glycine (mmol/l) | 0.14 (0.04) | 0.14 (0.04) |
| Glycoprotein acetyls (mmol/l) | 0.78 (0.11) | 0.81 (0.11) |
| Histidine (mmol/l) | 0.07 (0.01) | 0.06 (0.01) |
| Lactate (mmol/l) | 3.87 (1.06) | 3.84 (1.05) |
| Omega-3 fatty acids (mmol/l) | 0.49 (0.21) | 0.49 (0.21) |
| Phenylalanine (mmol/l) | 0.05 (0.01) | 0.05 (0.01) |
| Pyruvate (mmol/l) | 0.07 (0.03) | 0.08 (0.03) |
| Total branched-chain amino acids (mmol/l) | 0.37 (0.08) | 0.38 (0.08) |
| Total cholines (mmol/l) | 2.43 (0.36) | 2.45 (0.36) |
| Tyrosine (mmol/l) | 0.06 (0.01) | 0.06 (0.01) |
| Polygenic risk score | -0.03 (0.78) | 0.18 (0.81) |
| | **Prevalence of "yes" (%)** | |
| Antihypertensive medication | 2,880 (11.43) | 596 (22.34) |
| Atrial fibrillation | 211 (0.84) | 44 (1.65) |
| Atypical antipsychotic | 70 (0.28) | 8 (0.30) |
| Chronic kidney disease (stages 3-5) | 13 (0.05) | 0 (0.00) |
| Diabetes | 537 (2.13) | 116 (4.35) |
| Erectile dysfunction | 127 (0.50) | 29 (1.09) |
| Ethnicity: Black | 429 (1.70) | 24 (0.90) |
| Ethnicity: Other | 1,004 (3.98) | 94 (3.52) |
| Family history of coronary artery disease | 8,856 (35.14) | 1,189 (44.57) |
| Migraine | 369 (1.46) | 40 (1.50) |
| Rheumatoid arthritis | 164 (0.65) | 30 (1.12) |
| Severe mental health disorder | 108 (0.43) | 17 (0.64) |
| Smoker: current | 2,753 (10.92) | 392 (14.69) |
| Smoker: former | 8,993 (35.68) | 1,095 (41.04) |
| Systemic lupus erythematosus | 8 (0.03) | 3 (0.11) |
| Systemic steroid | 172 (0.68) | 31 (1.16) |

**(B)**

| Variable | Non-cases | Cases |
|---|---|---|
| *N* | 38,938 | 2,044 |
| | **Mean (s.d.)** | |
| Age (years) | 55.10 (7.95) | 60.00 (6.90) |
| Body mass index (kg/m2) | 26.61 (4.92) | 27.86 (5.39) |
| Systolic blood pressure: mean (mmHg) | 133.64 (18.80) | 142.10 (19.92) |
| Systolic blood pressure: s.d. (mmHg) | 5.37 (4.41) | 6.06 (4.90) |
| Townsend deprivation index | -1.46 (2.98) | -1.21 (3.10) |
| Alanine aminotransferase (U/L) | 19.46 (11.34) | 21.14 (13.99) |
| Albumin (g/L) | 44.96 (2.58) | 44.45 (2.63) |
| Alkaline phosphatase (U/L) | 83.11 (26.13) | 92.16 (28.07) |
| Apolipoprotein A1 (g/L) | 1.64 (0.27) | 1.60 (0.27) |
| Apolipoprotein B (g/L) | 1.05 (0.23) | 1.13 (0.24) |
| Aspartate aminotransferase (U/L) | 23.98 (8.58) | 25.47 (12.62) |
| Calcium (mmol/L) | 2.38 (0.10) | 2.39 (0.10) |
| Cholesterol (mmol/L) | 230.86 (41.69) | 241.86 (42.14) |
| C-reactive protein (mg/L) | 2.55 (4.17) | 3.61 (5.17) |
| Creatinine (µmol/L) | 63.96 (11.01) | 64.66 (11.75) |
| Cystatin C (mg/L) | 0.86 (0.14) | 0.93 (0.16) |
| Direct bilirubin (µmol/L) | 1.53 (0.66) | 1.44 (0.50) |
| Gamma glutamyltransferase (U/L) | 28.38 (29.29) | 34.00 (35.90) |
| Glucose (mmol/L) | 4.97 (0.82) | 5.12 (1.13) |
| Glycated hemoglobin (mmol/mol) | 34.93 (4.65) | 36.43 (6.04) |
| High-density lipoprotein cholesterol (mmol/L) | 62.38 (14.38) | 59.08 (13.96) |
| Insulin-like growth factor 1 (nmol/L) | 21.29 (5.67) | 19.77 (5.64) |
| Lipoprotein(a) (g/L) | 44.56 (48.76) | 48.55 (50.40) |
| Low density cholesterol (mmol/L) | 3.71 (0.84) | 3.98 (0.84) |
| Phosphate (mmol/L) | 1.19 (0.15) | 1.19 (0.15) |
| Sex hormone binding globulin (nmol/L) | 64.20 (31.44) | 61.62 (31.37) |
| Testosterone (nmol/L) | 1.11 (0.62) | 1.16 (0.77) |
| Total bilirubin (µmol/L) | 8.18 (3.73) | 7.75 (3.15) |
| Total protein (g/L) | 72.36 (4.08) | 72.43 (4.31) |
| Triglycerides (mmol/L) | 1.48 (0.81) | 1.75 (0.88) |
| Urate (µmol/L) | 264.79 (62.37) | 283.33 (68.64) |
| Urea (mmol/L) | 5.14 (1.23) | 5.41 (1.30) |
| Vitamin D (µg) | 48.47 (20.69) | 47.12 (19.96) |
| Basophil count (e9 cells/L) | 0.04 (0.05) | 0.04 (0.07) |
| Eosinophil count (e9 cells/L) | 0.16 (0.13) | 0.17 (0.12) |
| Hematocrit (%) | 39.14 (2.76) | 39.56 (2.78) |
| Hemoglobin concentration (g/dL) | 13.46 (0.95) | 13.61 (0.98) |
| High light scatter reticulocyte count (e12 cells/L) | 0.02 (0.01) | 0.02 (0.01) |
| Immature reticulocyte fraction | 0.29 (0.06) | 0.29 (0.06) |
| Lymphocyte count (e9 cells/L) | 1.99 (0.98) | 2.05 (0.66) |
| Mean corpuscular hemoglobin (pg) | 31.30 (1.88) | 31.29 (1.92) |
| Mean corpuscular hemoglobin concentration (g/dL) | 34.41 (0.98) | 34.41 (0.91) |
| Mean corpuscular volume (fL) | 90.92 (4.61) | 90.91 (4.95) |
| Mean platelet volume (fL) | 9.37 (1.09) | 9.33 (1.07) |
| Mean reticulocyte volume (fL) | 105.59 (7.68) | 105.82 (8.16) |
| Mean sphered cell volume (fL) | 83.08 (5.24) | 83.15 (5.66) |
| Monocyte count (e9 cells/L) | 0.43 (0.19) | 0.46 (0.21) |

| | | |
|---|---|---|
| Neutrophil count (e9 cells/L) | 4.14 (1.36) | 4.36 (1.48) |
| Nucleated red blood cell count (e9 cells/L) | 0.02 (0.05) | 0.02 (0.02) |
| Platelet count (e9 cells/L) | 266.32 (59.70) | 271.35 (61.10) |
| Platelet crit (%) | 0.25 (0.05) | 0.25 (0.05) |
| Platelet distribution width (%) | 16.41 (0.50) | 16.43 (0.49) |
| Red blood cell count (e12 cells/L) | 4.31 (0.33) | 4.36 (0.33) |
| Red blood cell distribution width (%) | 13.50 (1.04) | 13.60 (1.04) |
| Reticulocyte count (e9 cells/L) | 0.06 (0.03) | 0.06 (0.02) |
| White blood cell count (e9 cells/L) | 6.76 (1.90) | 7.08 (1.81) |
| 3-Hydroxybutyrate (mmol/l) | 0.06 (0.06) | 0.06 (0.06) |
| Acetate (mmol/l) | 0.02 (0.01) | 0.01 (0.02) |
| Acetoacetate (mmol/l) | 0.01 (0.01) | 0.01 (0.01) |
| Acetone (mmol/l) | 0.01 (0.01) | 0.01 (0.01) |
| Alanine (mmol/l) | 0.29 (0.07) | 0.29 (0.07) |
| Citrate (mmol/l) | 0.06 (0.01) | 0.06 (0.01) |
| Fatty acids: degree of unsaturation | 1.38 (0.07) | 1.37 (0.08) |
| Glutamine (mmol/l) | 0.52 (0.08) | 0.53 (0.08) |
| Glycine (mmol/l) | 0.19 (0.07) | 0.18 (0.07) |
| Glycoprotein acetyls (mmol/l) | 0.78 (0.12) | 0.83 (0.12) |
| Histidine (mmol/l) | 0.06 (0.01) | 0.06 (0.01) |
| Lactate (mmol/l) | 3.71 (1.07) | 3.73 (1.07) |
| Omega-3 fatty acids (mmol/l) | 0.55 (0.22) | 0.58 (0.23) |
| Phenylalanine (mmol/l) | 0.04 (0.01) | 0.05 (0.01) |
| Pyruvate (mmol/l) | 0.08 (0.03) | 0.08 (0.03) |
| Total branched-chain amino acids (mmol/l) | 0.33 (0.08) | 0.34 (0.08) |
| Total cholines (mmol/l) | 2.67 (0.39) | 2.71 (0.40) |
| Tyrosine (mmol/l) | 0.06 (0.01) | 0.06 (0.01) |
| Polygenic risk score | -0.00 (0.79) | 0.17 (0.82) |
| | **Prevalence of "yes" (%)** | |
| Antihypertensive medication | 4,504 (11.57) | 505 (24.71) |
| Atrial fibrillation | 131 (0.34) | 30 (1.47) |
| Atypical antipsychotic | 73 (0.19) | 3 (0.15) |
| Chronic kidney disease (stages 3-5) | 9 (0.02) | 1 (0.05) |
| Diabetes | 478 (1.23) | 71 (3.47) |
| Ethnicity: Black | 711 (1.83) | 26 (1.27) |
| Ethnicity: Other | 1,316 (3.38) | 59 (2.89) |
| Family history of coronary artery disease | 16,527 (42.44) | 1,121 (54.84) |
| Migraine | 1,784 (4.58) | 111 (5.43) |
| Rheumatoid arthritis | 505 (1.30) | 51 (2.50) |
| Severe mental health disorder | 161 (0.41) | 14 (0.68) |
| Smoker: current | 3,054 (7.84) | 253 (12.38) |
| Smoker: former | 12,353 (31.72) | 715 (34.98) |
| Systemic lupus erythematosus | 82 (0.21) | 10 (0.49) |
| Systemic steroid | 275 (0.71) | 42 (2.05) |

**Appendix Table 5.** Hazard ratios of covariates selected by LASSO stability selection in Cox models for cardiovascular disease incidence, fit on training data.

| | Men | Women |
|---|---|---|
| Age | 1.57 | 1.61 |
| *Albumin | 0.91 | 0.91 |
| Antihypertensive medication (Y/N) | 1.08 | 1.11 |
| *Apolipoprotein A1 | 0.9 | |
| *Apolipoprotein B | 1.17 | 1.08 |
| Atrial fibrillation (Y/N) | 1.03 | 1.06 |
| *C-reactive protein | 1.1 | 1.12 |
| Current smoking status | 1.09 | 1.2 |
| *Cystatin C | 1.1 | 1.2 |
| Family history of coronary artery disease (Y/N) | 1.14 | 1.13 |
| *Glycated hemoglobin | 1.06 | 1.04 |
| *Lipoprotein(a) | 1.12 | |
| Polygenic Risk Score | 1.22 | 1.17 |
| Systolic blood pressure: mean | 1.19 | 1.19 |
| Townsend deprivation index | 1.07 | |
| *Triglycerides | | 1.06 |
| *White blood cell count | 1.06 | |

*Variables are on log scale

**Appendix Table 6.** (A) Reclassification of cardiovascular disease cases and non-cases at 7.5% 10-year risk threshold with nested models, i.e., comparing pooled cohort equations (PCE) with Cox models using selected variables from LASSO stability selection alongside log hazards from PCE and (B) continuous and categorical net reclassification improvement (NRI) for these models and integrated discrimination improvement (IDI) compared with PCE, in men and women.

**A** Reclassification of cardiovascular disease cases and non-cases

Men

| PCE Predicted 10-year risk (%) | | LASSO stability selection: Predicted 10-year risk (%) <7.5 | LASSO stability selection: Predicted 10-year risk (%) ≥7.5 | Reclassified (%) |
|---|---|---|---|---|
| Cases | <7.5 | 871 | 376 | 30.2 |
| | ≥7.5 | 292 | 2,032 | 12.6 |
| Non-cases | <7.5 | 17,404 | 2,333 | 11.8 |
| | ≥7.5 | 3,151 | 10,060 | 23.9 |

Women

| PCE Predicted 10-year risk (%) | | LASSO stability selection: Predicted 10-year risk (%) <7.5 | LASSO stability selection: Predicted 10-year risk (%) ≥7.5 | Reclassified (%) |
|---|---|---|---|---|
| Cases | <7.5 | 1,733 | 346 | 16.6 |
| | ≥7.5 | 143 | 512 | 21.8 |
| Non-cases | <7.5 | 44,528 | 2,406 | 5.1 |
| | ≥7.5 | 1,815 | 3,309 | 35.4 |

**B** Continuous and categorical net reclassification improvement (NRI) and integrated discrimination improvement (IDI) for LASSO stability selection variables versus PCE

**Men**

| | Continuous NRI | Categorical NRI | IDI |
|---|---|---|---|
| **Cases** (N=3,571) | 0.121 (0.088; 0.153) | 0.024 (0.009; 0.038) | |
| **Non-cases** (N=32,948) | 0.197 (0.186; 0.208) | 0.025 (0.020; 0.029) | |
| **Full population** (N=36,519) | 0.318 (0.283; 0.352) | 0.048 (0.034; 0.063) | 0.0155 (0.0137; 0.0173) |

**Women**

| | Continuous NRI | Categorical NRI | IDI |
|---|---|---|---|
| **Cases** (N=2,734) | 0.120 (0.083; 0.157) | 0.074 (0.059; 0.090) | |
| **Non-cases** (N=52,058) | 0.224 (0.215; 0.232) | -0.011 (-0.014; -0.009) | |
| **Full population** (N=54,792) | 0.344 (0.305; 0.382) | 0.063 (0.047; 0.079) | 0.0094 (0.0081; 0.0108) |

**Appendix Figure 1.** Heatmap of Pearson's correlations between biochemistry (green), PCE/QRISK3 (blue), genetic (brown), hematology (red) and Nightingale (purple) markers. Correlations were evaluated separately in cases (N=4,712) and controls (N=64,143) with complete data. Measured levels of biochemistry, hematology and Nightingale biomarkers were log-transformed.

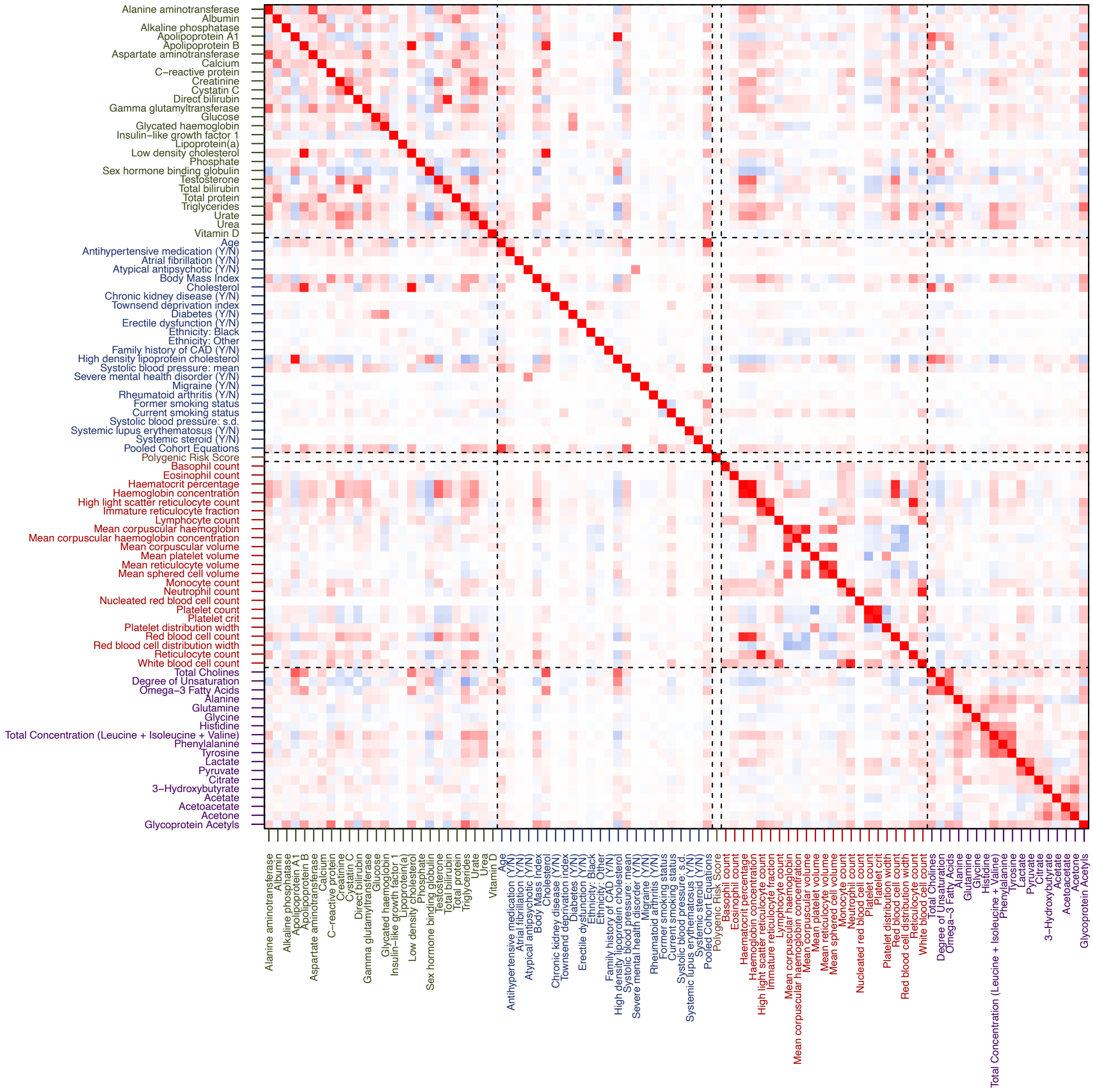

**Appendix Figure 2.** Calibration plots showing predicted and observed 10-year risk of cardiovascular disease by decile of log hazard using pooled cohort equations in men (A-D) and women (E-H). Probabilities computed in training (A, E: non-recalibrated; B, F: recalibrated) and test sets (C, G: non-recalibrated; D, H: recalibrated). Pooled cohort equations recalibrated by using computed log hazards as predictor in Cox models fitted on sex-stratified training sets. Slope (β) are reported.

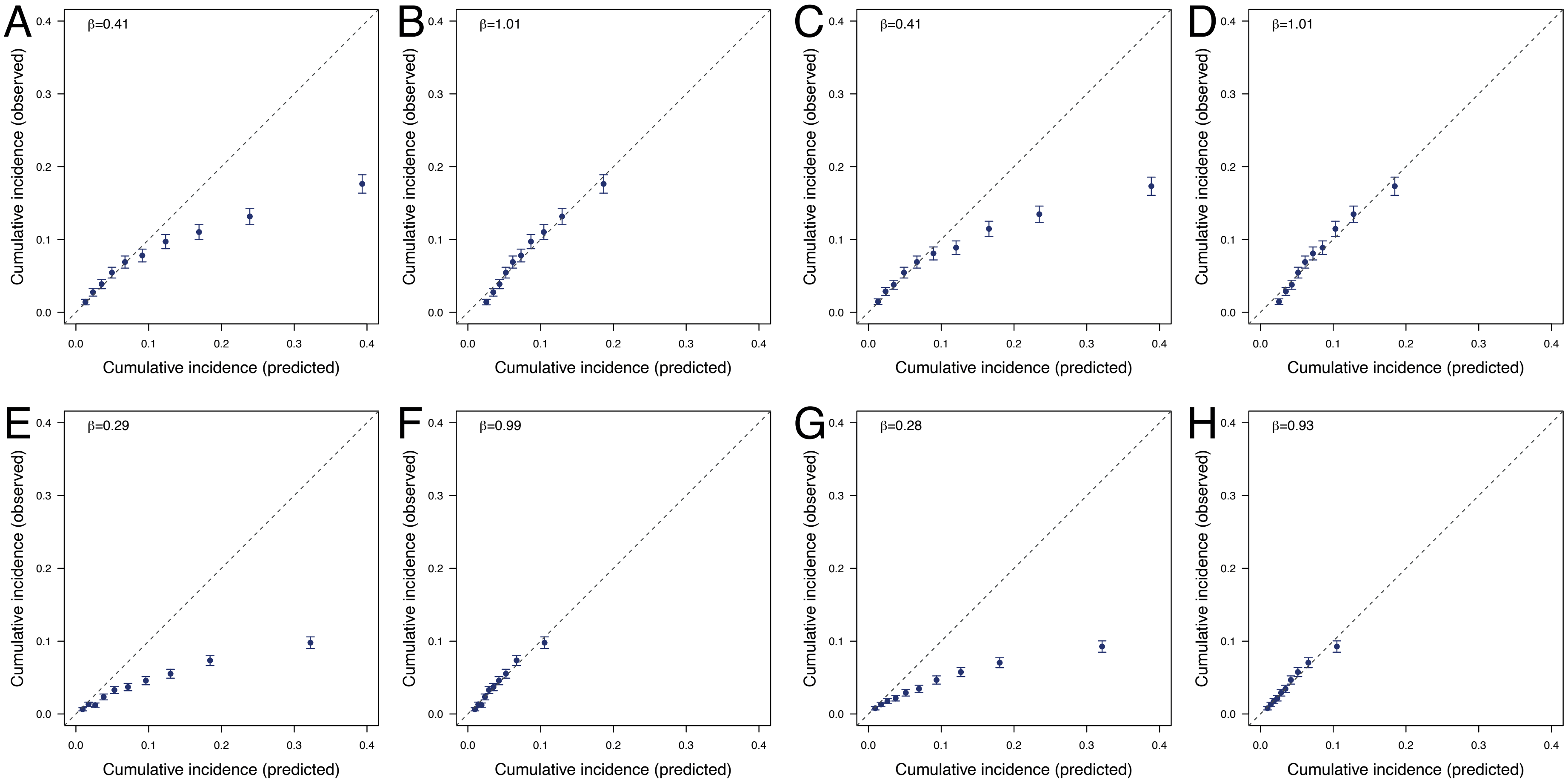

**Appendix Figure 3.** C-statistics in test data using Cox models sequentially including predictors in descending order of selection proportion in men (A) and women (B). Results are presented for all variables, and the vertical red dashed line indicates the model including all stably selected variables. C-statistics for recalibrated PCE are also reported (black horizontal dashed line). Color code: Variables in either PCE or QRISK3 – blue; biochemistry – green; hematology – red; polygenic risk score – brown.

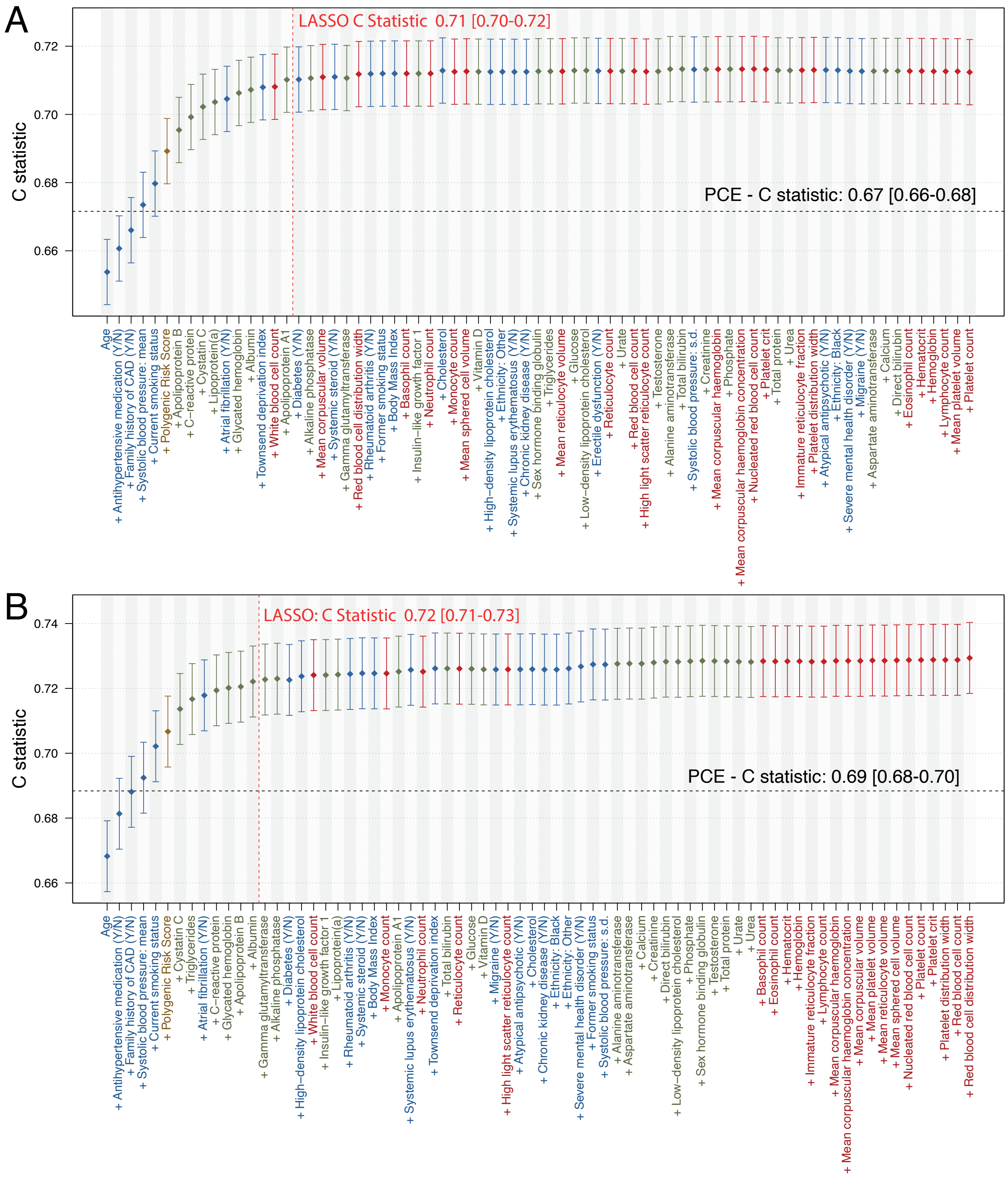

**Appendix Figure 4.** Scatter plots comparing the LASSO stability selection proportions from the base model and model where pooled cohort equations (PCE) log hazards are included in place of their constituent variables. Results are reported in (A) men and (B) women separately.

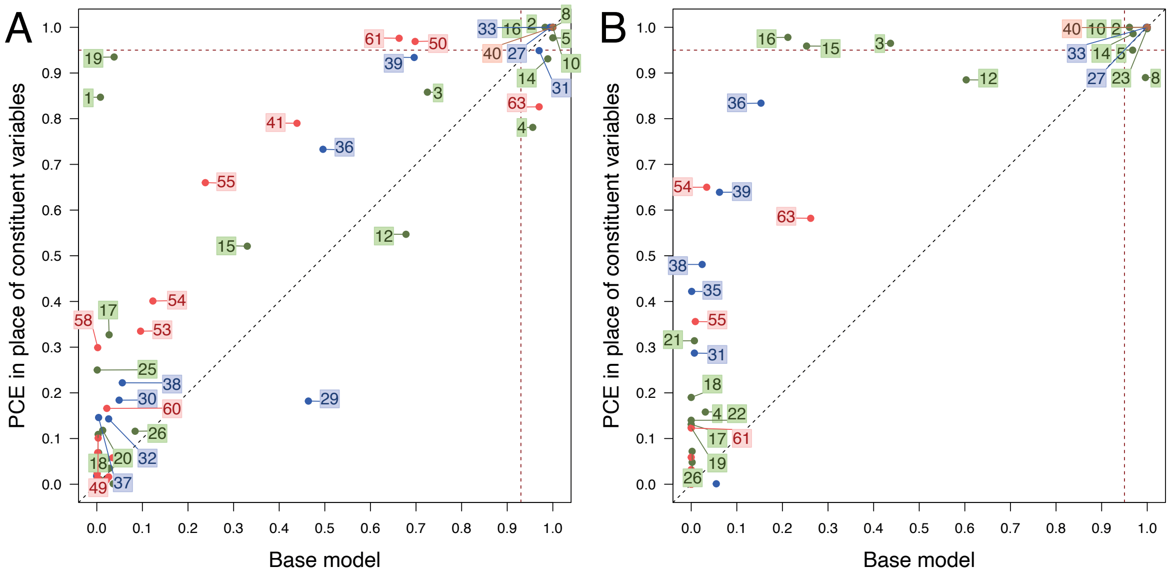

1 – Alanine aminotransferase
2 – Albumin
3 – Alkaline phosphatase
4 – Apolipoprotein A1
5 – Apolipoprotein B
6 – Aspartate aminotransferase
7 – Calcium
8 – C–reactive protein
9 – Creatinine
10 – Cystatin C
11 – Direct bilirubin
12 – Gamma glutamyltransferase
13 – Glucose
14 – Glycated hemoglobin
15 – Insulin–like growth factor 1
16 – Lipoprotein(a)
17 – Low density cholesterol
18 – Phosphate
19 – Sex hormone binding globulin
20 – Testosterone
21 – Total bilirubin
22 – Total protein
23 – Triglycerides
24 – Urate
25 – Urea
26 – Vitamin D
27 – Atrial fibrillation (Y/N)
28 – Atypical antipsychotic (Y/N)
29 – Body Mass Index
30 – Chronic kidney disease (Y/N)
31 – Townsend deprivation index
32 – Erectile dysfunction (Y/N)
33 – Family history of CAD (Y/N)
34 – Severe mental health disorder (Y/N)
35 – Migraine (Y/N)
36 – Rheumatoid arthritis (Y/N)
37 – Systolic blood pressure: s.d.
38 – Systemic lupus erythematosus (Y/N)
39 – Systemic steroid (Y/N)
40 – Polygenic Risk Score
41 – Basophil count
42 – Eosinophil count
43 – Hematocrit percentage
44 – Hemoglobin concentration
45 – High light scatter reticulocyte count
46 – Immature reticulocyte fraction
47 – Lymphocyte count
48 – Mean corpuscular haemoglobin
49 – Mean corpuscular haemoglobin concentration
50 – Mean corpuscular volume
51 – Mean platelet volume
52 – Mean reticulocyte volume
53 – Mean sphered cell volume
54 – Monocyte count
55 – Neutrophil count
56 – Nucleated red blood cell count
57 – Platelet count
58 – Platelet crit
59 – Platelet distribution width
60 – Red blood cell count
61 – Red blood cell distribution width
62 – Reticulocyte count
63 – White blood cell count
64 – Alanine aminotransferase

**Appendix Figure 5.** C statistics in test data using Cox models sequentially including predictors in descending of selection proportion from the base model (A, B in men and women, respectively) and the model further including metabolomic variables (C, D in men and women, respectively). Results are presented for all variables, and the vertical red dashed line indicates the model including all stably selected variables. C-statistics for recalibrated PCE are also reported (black horizontal dashed line). Color code: Variables in either PCE or QRISK3 – blue; biochemistry – green; hematology – red; polygenic risk score – brown.

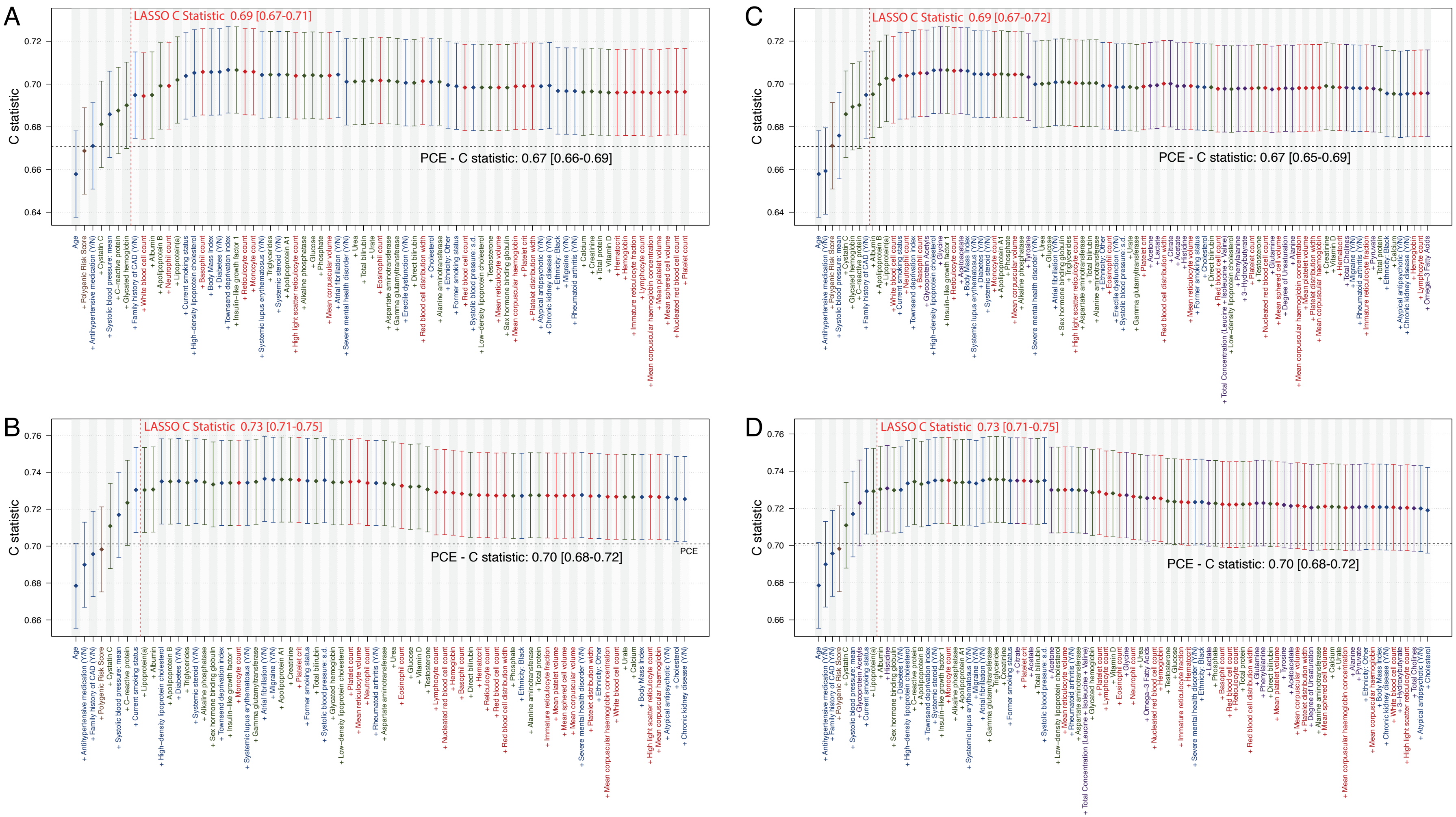

**Appendix Figure 6** Receiver operating characteristic (ROC) curves from logistic models for incident CVD, excluding (top panel: A and B in men and women, respectively) and including (bottom panel: C and D in men and women, respectively) NMR-derived metabolomic variables. Results from logistic models including PCE constituents (blue line) or sex-specific LASSO stably selected variables (red line) are shown, with mean area under the curve (AUC) and 95% confidence intervals.

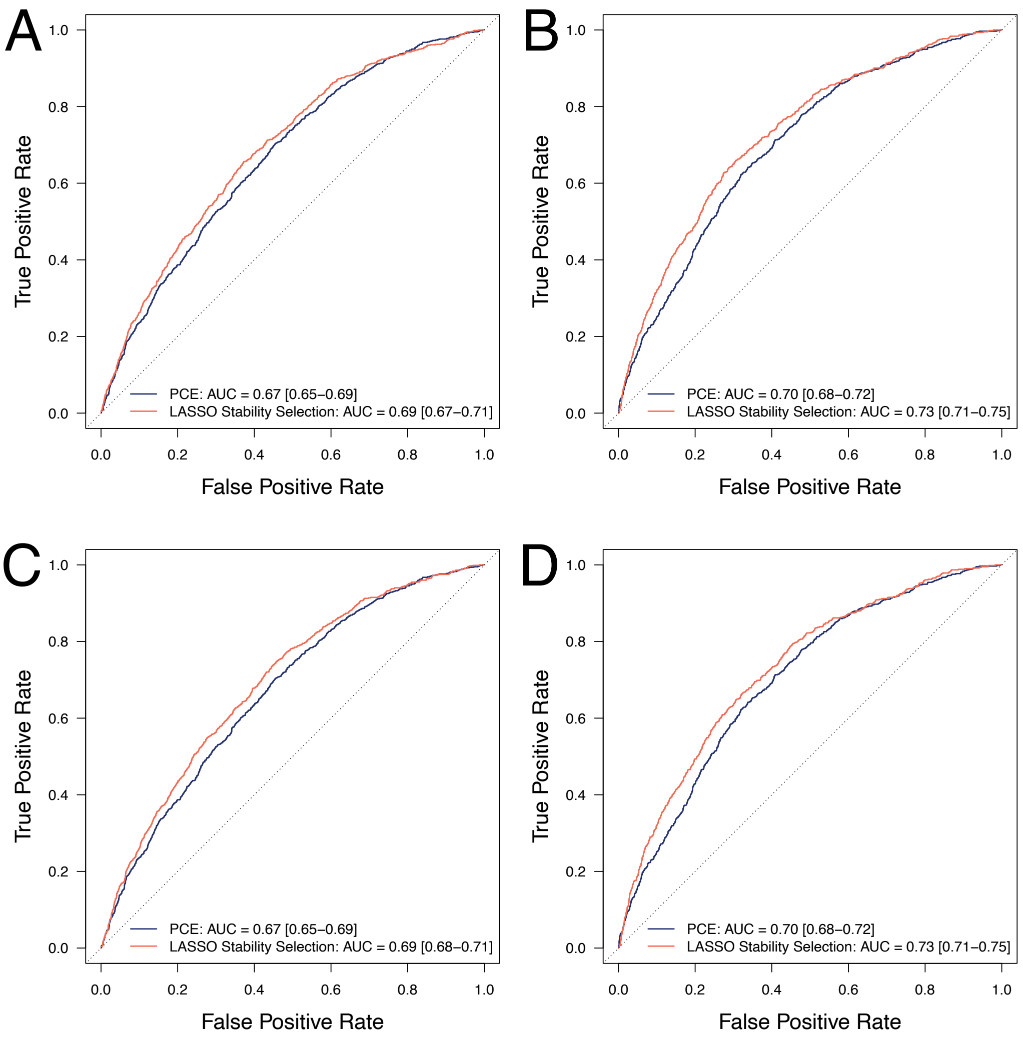
